# Differentially expressed genes reflect disease-induced rather than disease-causing changes in the transcriptome

**DOI:** 10.1101/2020.10.19.20213538

**Authors:** Eleonora Porcu, Marie C. Sadler, Kaido Lepik, Chiara Auwerx, Andrew R. Wood, Antoine Weihs, Diogo M. Ribeiro, Stefania Bandinelli, Toshiko Tanaka, Matthias Nauck, Uwe Völker, Olivier Delaneau, Andres Metspalu, Alexander Teumer, Timothy Frayling, Federico A. Santoni, Alexandre Reymond, Zoltán Kutalik

**Author notes:** corresponding author: Eleonora Porcu.

## Abstract

Comparing transcript levels between healthy and diseased individuals allows the identification of differentially expressed genes, which may be causes, consequences or mere correlates of the disease under scrutiny. Here, we propose a bi-directional Transcriptome-Wide Mendelian Randomization (TWMR) approach that integrates summary-level data from GWAS and whole-blood eQTLs in a MR framework to investigate the causal effects between gene expression and complex traits.

Whereas we have previously developed a TWMR approach to elucidate gene expression to trait causal effects, here we are adapting the method to shed light on the causal imprint of complex traits on transcript levels. We termed this new approach reverse TWMR (revTWMR). Integrating bi-directional causal effects between gene expression and complex traits enables to evaluate their respective contributions to the correlation between gene expression and traits. We uncovered that whole blood gene expression-trait correlation is mainly driven by causal effect from the phenotype on the expression rather than the reverse. For example, BMI- and triglycerides-gene expression correlation coefficients robustly correlate with trait-to-expression causal effects (*r*=0.09, *P*=1.54×10^−39^ and *r*=0.09, *P*=1.19×10^−34^, respectively), but not detectably with expression-to-trait effects.

Genes implicated by revTWMR confirmed known associations, such as rheumathoid arthritis and Crohn’s disease induced changes in expression of *TRBV* and *GBP2*, respectively. They also shed light on how clinical biomarkers can influence their own levels. For instance, we observed that high levels of high-density lipoprotein (HDL) cholesterol lowers the expression of genes involved in cholesterol biosynthesis (*SQLE, FDFT1*) and increases the expression of genes responsible for cholesterol efflux (*ABCA1, ABCG1*), two key molecular pathways in determining HDL levels. Importantly, revTWMR is more robust to pleiotropy than polygenic risk score (PRS) approaches which can be misled by pleiotropic outliers. As one example, revTWMR revealed that the previously reported association between educational attainment PRS and *STX1B* is exclusively driven by a highly pleiotropic SNP (rs2456973), which is strongly associated with several hematological and anthropometric traits.

In conclusion, our method disentangles the relationship between gene expression and phenotypes and reveals that complex traits have more pronounced impact on gene expression than the reverse. We demonstrated that studies comparing the transcriptome of diseased and healthy subjects are more prone to reveal disease-induced gene expression changes rather than disease causing ones.

## Introduction

To unravel the genetics of complex diseases and traits causes, multiple approaches have concentrated on gene expression, in particular by contrasting the expression of RNA transcripts in two different groups of samples to understand how genes are expressed in health and disease [1-4]. The purpose of such an approach is to identify differentially expressed genes (DEG) that can be used to obtain mechanistic insights from diseases or serve as clinical biomarkers for early diagnostics. However, DEG analyses suffer from the inability to distinguish between causes, consequences or mere correlations between gene expression and phenotypes. Ideally, to understand the contributions to the observed trait-expression correlations, both the assessment of bi-directional causal effects and the impact of (unmeasured) confounders is needed. In turn we argue that if the observed correlations and bi-directional causal effects are estimated, the contribution of such confounders can be gauged.

Genome-wide association studies (GWAS) identified thousands of common genetic variants associated with complex human traits [5] and studies on expression quantitative trait loci (eQTLs) showed how genetic variants contribute to the regulation of gene expression levels [6]. The overlay of the two methodologies showed that trait-associated SNPs are three times more likely to be eQTLs [7-10], suggesting that gene expression is a reliable intermediary between DNA variation and higher-order complex phenotypes. Starting from this hypothesis, many statistical approaches integrating GWAS and eQTLs summary statistics have been proposed to detect these overlapping associations [9, 11, 12]. However, while these studies aim to identify genes whose (genetically determined) expression is significantly associated to complex traits, they do not aim to estimate the strength of the causal effect and are unable to distinguish causation from pleiotropy (i.e. when a genetic variant independently affects gene expression and phenotype). The most efficient way to address this challenge is to combine summary level data from eQTL and GWAS studies in a two-sample Mendelian Randomization framework [13] to evaluate whether gene expression has a causal influence on a complex trait. Such methods successfully identified thousands of genes associated with complex traits.

Yet, these transcriptome-wide approaches use only *cis*-eQTLs as instruments to tease out the causal effect of gene expression on a complex trait while the variation in gene expression may be secondary to, rather than causal for, the disease process (‘reverse causation’). Disease-associated genetic variants affect expression levels more often in *trans* than in *cis* [14]. Hence, polygenic risk scores (PRS) have been used to evaluate the association between genetically predicted complex traits and gene expression levels [14]. However, such PRS-based approaches are prone to detect mere associations due to pleiotropic SNPs. To circumvent this issue and elucidate the impact of diseases on the transcriptome program at a large scale in a principled way, we applied a reverse Transcriptome-Wide Mendelian Randomization approach (called revTWMR), which integrates summary-level data from GWAS and *trans*-eQTLs studies in a MR framework to estimate the causal effect of phenotypes on gene expression.

By combining revTWMR results with the causal effects of gene expression on phenotypes - estimated by Transcriptome-Wide Mendelian Randomization (TWMR) [15] - we obtained a clear picture of the bi-directional causal effects between gene expression and complex traits (Figure 1) and evaluated their contribution to their observational correlation.

**Figure 1.**
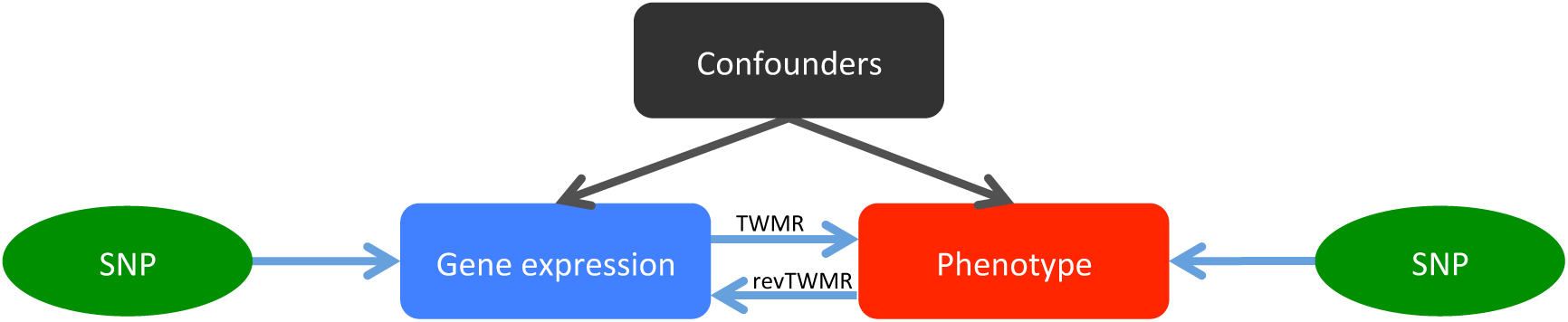
Schematic representation of how TWMR and revTWMR dissect bi-directional causal and confounder contributions to the observed correlation between gene expression and phenotype.

## Results

### Overview of the approach

We have recently developed a transcriptome-wide summary statistics-based Mendelian Randomization approach (TWMR [15]) integrating summary-level data from GWAS and *cis*-eQTL studies. Applying TWMR to summary data from whole blood *cis*-eQTL meta-analyses from > 32,000 individuals (eQTLGen Consortium [14]) and to the largest publicly available GWAS summary statistics revealed an atlas of putative functionally relevant genes for several complex human traits [15]. This approach can be reversed to design a multi-instrument MR approach to estimate the causal effect of a phenotype (exposure) on gene expression (outcome) (revTWMR, Figure 1). For each gene, using the inverse–variance weighted meta-analysis of ratio estimates from summary statistics [16], we estimate the causal effect of a phenotype on the expression of the probed gene as

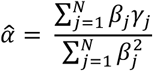

where *β*_*j*_ and *γ*_*j*_ are the effect sizes of *SNP*_*j*_ on the phenotype and on the expression level of the probed gene, respectively, and *N* is the number of independent SNPs used as instrument variables.

### Applying revTWMR to GWAS and eQTL summary statistics

We applied revTWMR to data to assess causal associations between 12 complex traits - body mass index (BMI), Crohn’s disease (CD), educational attainment (EDU), fasting glucose (FG), high-density lipoprotein (HDL), height, low-density lipoprotein (LDL), rheumatoid arthritis (RA), schizophrenia (SCZ), total cholesterol (TC), triglycerides (TG) and waist-to-hip ratio adjusted for BMI (WHRadjBMI) - and the expression of 19,942 genes. We combined summary whole blood *trans*-eQTLs data from the eQTLGen Consortium [14], with large publicly available GWAS for the traits of interest [17-25]. In parallel, we performed TWMR analyses on the same set of traits, allowing to test for the presence of bi-directional effects. In total, we found 69 genes significantly affected by at least one phenotype, often corroborating known biological associations (*P*_revTWMR_ < 2.5 × 10^−6^ = 0.05/19,942) (Supplementary Table 1).

**Table 1.** Correlation between observational phenotype-gene expression correlation and revTWMR and TWMR effects. For each phenotype available in at least two cohorts, we calculated the correlation between the observational correlation estimates and the revTWMR and TWMR effects. For significant correlations, we computed the adjusted correlation correcting for regression dilution bias.

| Trait | revTWMR |  | TWMR |  |
| --- | --- | --- | --- | --- |
|  | correlation (adjusted) | Pvalue | correlation | Pvalue |
| BMI | 0.09 (0.46) | 1.54E-39 | 0 | 0.56 |
| Educational attainment | 0.05 (0.27) | 1.30E-12 | 0.01 | 0.11 |
| Fasting Glucose | 0.08 (0.57) | 5.58E-17 | 0.02 | 0.82 |
| HDL | 0.06 (0.22) | 1.35E-15 | 0 | 0.56 |
| height | 0.11 (0.72) | 1.95E-53 | 0.02 | 0.01 |
| LDL | 0.01 (0.06) | 0.24 | 0.01 | 0.39 |
| Total Cholesterol | 0.02 (0.25) | 5.44E-04 | 0.01 | 0.27 |
| Triglycerides | 0.09 (0.26) | 1.19E-34 | 0 | 0.57 |
| Waist-to-hip ratio | 0.04 (0.51) | 1.14E-07 | 0 | 0.55 |

The most influential trait in our analysis was rheumathoid arthritis, which significantly influenced the expression of 33 genes. These were analyzed for functional enrichment with KEGG pathway [26], Gene Ontology [27] and InterPro [28]. Immunoglobulin InterPro functional groups (IPR013106, IPR007110 and IPR013783) were the top significantly enriched classes (Supplementary Table 2). Closer investigation revealed that this enrichment is due to the presence of 11 T cell receptor α and β variable genes (*TRAV* and *TRBV*). Interestingly, a bias in Vβ gene utilization by T cells in patients suffering from RA was reported [29]. In addition, many other immune related genes were identified. For instance, RA positively increases the expression of *GZMK* (*α*_revTWMR_ = 0.11, *P*_revTWMR_ = 5.2×10^−07^), a protease that confers cytolytic activity to natural killer cells and T lymphocytes [30]. Accordingly, *GZMK*^*+*^ CD8 T cells were found to induce inflammation in the synovium of RA patients [31] and elevated expression of *GZMK* was proposed as a biomarker to discern RA patients from those suffering from the related osteoarthritis [32]. Our analysis also revealed genes influenced by Crohn’s disease, another autoimmune disease. Guanylate-binding protein (GBP) levels, which are increased in individuals with inflammatory bowel diseases, are considered as a marker interferon γ-activated cells [33]. We confirmed the association by observing increased expression of *GBP2* (*α*_revTWMR_ = 0.079, *P*_revTWMR_ = 1.3×10^−08^) in CD patients. We similarly confirmed the role of *STAT1* in CD (*α*_revTWMR_ = 0.075, *P*_revTWMR_ = 1.4×10^−07^): the positive causal effect reflects the previously reported up-regulation of the gene in CD patients [34].

As the other traits influenced a smaller number of genes, no further significant enrichments were found. Nevertheless, a gene-by-gene investigation revealed many known or highly plausible associations. For instance, we found a significant effect of body mass index (BMI) on *ALDH1A1* (*α*_revTWMR_ = −0.26, *P*_revTWMR_ = 2.3×10^−08^), an enzyme that converts retinaldehyde to retinoic acid [35]. Retinoids have long been implicated in adipogenesis [36, 37] and *ALDH1A1* expression in visceral adipose tissue was shown to positively correlate with BMI [38]. Focusing on genes affected by serum lipid levels (Supplementary Table 3), revTWMR revealed 13 genes whose expression is altered by HDL cholesterol and 20 by TG levels. In line with the commonly observed negative correlation between HDL and TG [39], 9 of these genes were impacted by both traits with an opposite direction of the causal effect (Supplementary Table 1). Regarding the impact of high HDL levels, we found that it reduces the expression of squalene synthase (*FDFT1*; *α*_revTWMR_ = −0.17, *P*_revTWMR_ = 2.3×10^−10^) and squalene epoxidase (*SQLE*; *α*_revTWMR_ = −0.15, *P*_revTWMR_ = 2.7×10^−09^), two key enzymes of the cholesterol biosynthesis pathway [40, 41]. Interestingly, serum levels of squalene, the product and substrate of squalene synthase and epoxidase, respectively, negatively correlate with HDL-cholesterol [42]. According to revTWMR, genes involved in cholesterol transport were impacted too: high HDL negatively impacts the expression of the LDR receptor (*LDLR*; *α*_revTWMR_ = −0.16, *P*_revTWMR_ = 2.8×10^−10^), while having a positive impact on *MYLIP* (also known as *IDOL*; *α*_revTWMR_ = 0.27, *P*_revTWMR_ =1.4×10^−27^), a ubiquitin ligase that induces degradation of the LDL receptor [43]. In parallel, HDL seems to increase the expression of the *ABCA1* (*α*_revTWMR_ = 0.29, *P*_revTWMR_ =5.5×10^−30^) and *ABCG1* (*α*_revTWMR_ = 0.27, *P*_revTWMR_ =7.1×10^−28^), two transporters responsible for cholesterol efflux from macrophages [44]. *ABCA1* was also the only gene impacted by total cholesterol levels (TC; *α*_revTWMR_ = −0.17, *P*_revTWMR_ =5.8×10^−08^). While we did not observe a significant effect of *ABCA1* on HDL and TC levels through TWMR, a GWAS has previously reported an association between *ABCA1* and these two traits [24], suggesting a complex regulatory mechanism. Together, these results are reminiscent of the well-described negative feedback mechanisms that tightly control cholesterol biosynthesis and uptake [45].

Despite strong indications of functional relevance, most revTWMR-implicated genes fall into genomic regions completely missed by GWAS, as is illustrated by the fact that revTWMR p-values are completely uncorrelated (*r* < 0.05) with those obtained by classical gene-based GWAS test performed using PASCAL [46] (See Methods; Supplementary Figure 1). In line with this observation, only two out of the 69 revTWMR-identified genes were significant for TWMR: *AFF3* and *FADS1*, known susceptibility loci for RA [47] and HDL levels [24], respectively, show a negative (*α*_*TWMR*_ = 0.08, *P*_TWMR_ = 1.6×10^−04^ and *α*_revTWMR_ = −0.13, *P*_revTWMR_ = 3.0×10^−07^) and positive (*α*_*TWMR*_ = −0.06, *P*_TWMR_ = 7.8×10^−11^ and *α* _revTWMR_ = −0.12, *P*_revTWMR_ = 4.8×10^−07^) causal feedback loop between the traits, respectively. The complexity of the *FADS1*-HDL interaction was recently highlighted by Dumont et al., who suggested that dietary intake of linoleic acid can modulate the impact of rs174547, a *FADS1* intronic variant, on HDL-cholesterol levels [48].

### Pleiotropic SNPs lead to biased causal effect estimates

The validity of revTWMR, as any MR approach, relies on three assumptions about the instruments: (i) they must be sufficiently strongly associated with the exposure; (ii) they should not be associated with any confounder of the exposure-outcome relationship; and (iii) they should be associated with the outcome only through the exposure. The third assumption (no pleiotropy) is crucial as MR causal estimates will be biased if the genetic variants have pleiotropic effects [49, 50]. Accordingly, revTWMR assumes that all genetic variants used as instrumental variables affect the gene expression only through the phenotype under scrutiny and not through independent biological pathways.

To test for the presence of pleiotropy, we used a similar approach to MR-PRESSO global test [50, 51], performing Cochran’s Q test. Under the assumption that the majority of SNPs influence gene expression only through the phenotype tested in the model, SNPs violating the third MR assumption would significantly increase the Cochran’s heterogeneity Q statistic (see Methods) and hence can be detected and removed. This was the case for 65 of the 119 originally significant trait → gene associations. Out of these 65 associations, 21 passed the heterogeneity test after removing up to 5 pleiotropic SNPs from the instrumental variables. Moreover, this procedure led to the identification of 4 additional associations initially masked by heterogeneity, giving the final number of 79 robust associations (Supplementary Table 1). Importantly, this method discriminates likely causal effects from pleiotropy. This is well illustrated by the example of *STX1B*, a gene that was found to be associated with educational attainment (EDU) through a PRS approach (*P*_PRS_ = 1.25×10^−20^) [14]. While revTWMR originally detected a significant association between EDU and *STX1B*, the association failed the heterogeneity test. After excluding the highly pleiotropic variant rs2456973 (strongly associated with hematological and anthropometric traits [52], Supplementary Table 4) from the instruments, the effect of EDU on *STX1B* was lost (Supplementary Figure 2).

**Figure 2.**
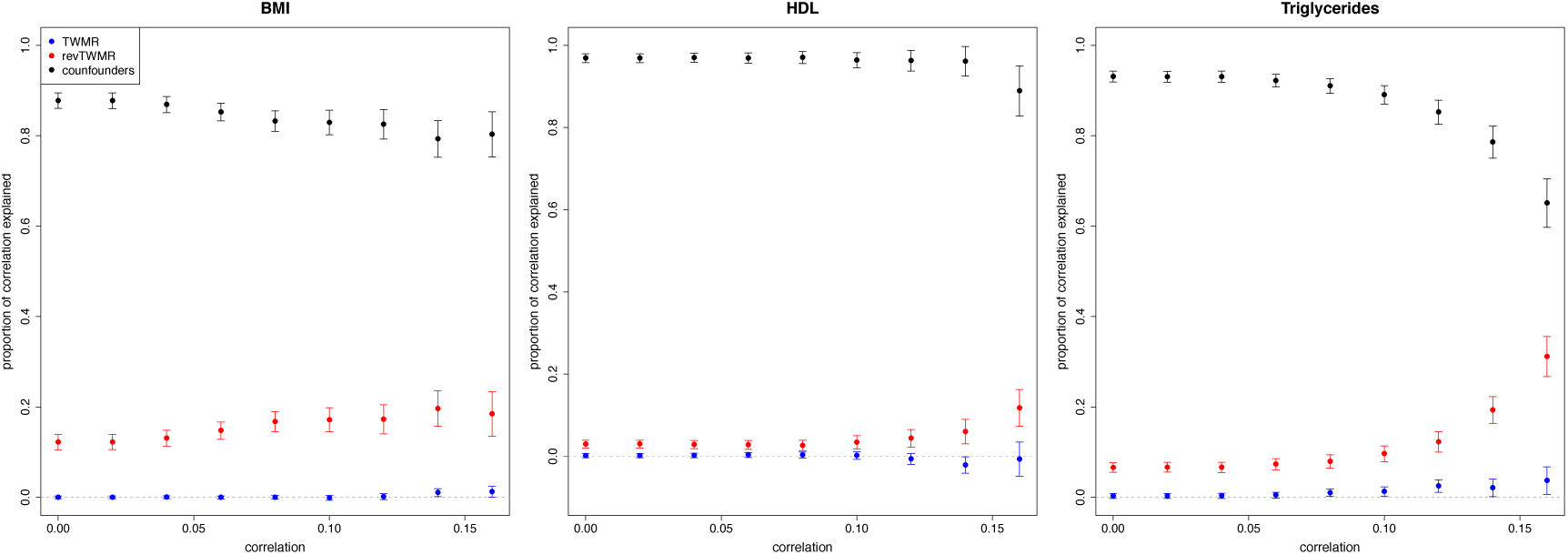
Partitioning of the gene expression-trait observational correlation for BMI, HDL and triglycerides. For each bin of correlation (absolute value) we plotted the combined contributions of the forward (TWMR, blue dots) and reverse (revTWMR, red dots) effect of the gene expression on the trait, the contribution of confounders (black dots) and their confidence interval.

### Trait correlation

Exploring the shared effect of complex traits and diseases on the transcriptional programs can provide useful etiological insights. Hence, for every phenotype-pair (*P*_*i*_, *P*_*j*_) we computed the gene expression perturbation correlation 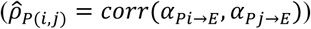 between the respective causal effect estimates of each phenotype on the gene expression (i.e. the Z scores from our revTWMR analysis) across a subset of 2,974 independent genes across the genome [15]. Among the 66 pairs of traits, we found 21 significant correlations (FDR<1%). We compared these results with the genetic correlation 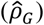 between traits estimated by LD score regression [53] and found a remarkable concordance between the two estimates (*r* = 0.80). Of note, 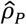 seems to be 55% of 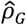 on average. Although 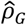 having smaller variance may explain part of this attenuation, we think that the main reason behind this observation is that only a part of 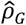 translates into consequences on gene–expression level in whole blood (Supplementary Figure 3). In particular, 8 pairs of traits showed significance for both 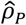 and 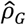, whereas 12 were significant only for 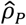, and 8 only for 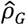. Among the significant correlations not identified by LD score regression 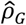, we found that HDL and LDL are negatively correlated 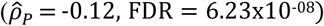 and that RA positively correlated with several traits: Crohn’s disease 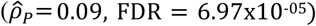, schizophrenia 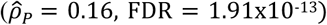, height 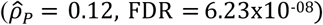, total cholesterol 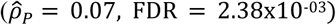 and triglycerides 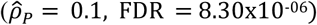 (Supplementary Table 5).

### Observational correlation is driven by reverse association

As a proof-of-concept, we asked how highly revTWMR-identified causal genes would rank in a DEG analysis. To address this question, we collected the observational correlation estimates between whole blood gene expression levels and the quantitative traits in three independent European cohorts (EGCUT (N = 488), InChianti (N = 609), and SHIP-Trend (N = 991)).

Correlating revTWMR effects to observational correlations (equivalent to DEG analysis), we found a significant agreement for all the traits, with the exception of LDL, for which we speculate that the lack of agreement might be due to the low number of significantly correlated genes, as well as the lack of concordance among cohorts (Table 1, Supplementary Figure 4). We re-estimated these correlations accounting for the error in the compared estimates (regression dilution bias) (see Methods). No significant correlation between observational correlations and the causal effects of the gene expression on phenotypes estimated by TWMR was observed (Table 1). Of note, when we correlated the P-values of the observational correlations with those obtained by conventional gene-based tests using GWAS results, we detected a significant concordance only for triglycerides (*r* = 0.02, *P* = 3.50×10^−04^) (Supplementary Table 6).

As TWMR and revTWMR results showed that causal feedback loops are rare (i.e. *α*_*TWMR*_ * *α*_*revTWMR*_ = 0), the observational correlation (*r*) can be approximated as the sum of the bi-directional effects estimated by TWMR and revTWMR plus the contribution of the confounding factors (see Methods). Hence we were able to calculate the proportion of correlation due to confounders. For each gene we calculated the contribution of TWMR and revTWMR as 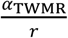 and 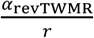, respectively. Consequently, the contribution of confounders is 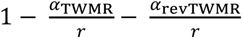. In each correlation bin (Figure 2) we combined such contributions using inverse variance meta-analysis and revealed that the observed correlation between gene expression and phenotype is mainly driven by confounders. For example, for genes correlated (|*r*| > 0.1) with BMI, 83% (*P* < 5.0×10^−324^) of the correlation is due to the confounders, 17% (*P* = 1.12×10^−36^) to the effect of BMI on gene expression and 0% (*P* = 0.65) to the forward effect (Figure 2 and Supplementary Table 7). We observed the same scenario for triglycerides: 89% (*P* < 5.0×10^−324^) of the correlation is due to confounders and 10% (*P* = 1.02×10^−26^) and 1% (*P* = 0.012) are due to reverse and forward effect of the gene expression on height respectively. For HDL we observed a stronger effect due to confounders (96%, *P* < 5.0×10^−324^) and a mild reverse effect contribution (4%, *P* = 2.40×10^−05^) (Figure 2).

### Genes affected by lipid traits are linked to drug targets

We assessed whether the protein products of the transcripts perturbed by a disease/trait in our revTWMR analysis are targets of drugs used to treat the disease in question. We started by defining a set of drugs relevant to the traits under investigation according to DrugBank [13]. Next, we retrieved high confidence interactions (confidence score > 0.7) involving these drugs, from STITCH, a manually curated database of predicted and experimental chemical-protein interactions [12]. We then searched for proteins that were a) identified as dysregulated by revTWMR and b) targeted by a drug indicated for the treatment of a given trait.

The gene product of four out of the 13 genes detected by revTWMR for low HDL met these criteria: phospholipid-transporting ATPase ABCA1 (*ABCA1*), squalene synthase (*FDFT1*), low-density lipoprotein receptor (*LDLR*) and sterol regulatory element-binding protein 1 (*SREBF1*) which interact with atorvastatin, lovastatin, pravastatin and simvastatin. We found that these four genes are also dysregulated by high triglyceride levels and *ABCA1* was the only detected gene dysregulated by high total cholesterol. We did not find drug targets among the genes significantly dysregulated by Crohn’s disease, rheumatoid arthritis and schizophrenia (Supplementary Table 8).

### Tissue-specific effects

As many traits manifest themselves only in certain tissues, it is essential to integrate data from the tissue of interest to estimate the impact of a phenotype at the transcriptome level. For this reason, we performed tissue-specific revTWMR analyses using tissue-specific *trans*-eQTLs identified by the Genotype Tissue Expression Project (GTEx) [54], which provides a unified view of genetic effects on gene expression across 49 human tissues. We tested the 79 previously identified significant trait → gene associations found in whole blood and detected three genes showing tissue-specific associations (*P*_revTYMR_ < 0.05/79, Supplementary Table 9). We found a negative effect of RA on *MYO1B* in the kidney cortex (*α*_revTWMR_ = −1.71, *P*_revTWMR_ = 2.4×10^−04^), as well as a significant effect of HDL on *LDLR* (*α*_revTWMR_ = −0.63, *P*_revTWMR_ = 4.3×10^−04^) and *ABCG1* (*α*_revTWMR_ = 0.61, *P*_revTWMR_ = 6.1×10^−04^) in the thyroid, a key tissue for lipid metabolism regulation [55]. Of note, the effect sizes of these associations are >6 fold larger than those estimated using whole blood data.

## Discussion

We presented a Mendelian randomization approach to study the impact of human phenotypes on the transcriptome. Across the 69 genes identified by our revTWMR approach, we observed a clear trend for functional relevance. Genes perturbed by complex diseases seem to confirm several previously reported associations between immune-related genes and autoimmune diseases such as RA (*TRBV, GZMK*) [29, 31, 32] and CD (*GBP2, STAT1*) [33, 34]. In addition, revTWMR could be used as a tool to gain insight into the regulatory mechanisms controlling biological pathways, as illustrated with our example regarding serum lipid levels. We observed that high HDL-cholesterol lowers the expression of genes involved in cholesterol biosynthesis (*SQLE, FDFT1*) and cellular cholesterol uptake (*LDLR*), while it increases the expression of genes responsible for degradation of *LDLR* (*MYLIP*) and cholesterol efflux (*ABCA1, ABCG1*). Together, this suggests that high HDL levels prevent intracellular cholesterol overload, which could explain its known cardioprotective effects [56]. However, TG levels, which were shown to independently increase risk of coronary artery disease (CAD) [57], impact the expression of the same genes in the opposite direction. An alternative scenario could be that high TG levels increase CAD risk through intracellular accumulation of cholesterol. The biological relevance of our findings is further supported by our drug target analysis, which found that four genes (*SREBF1, FDFT1, LDLR* and *ABCA1*) whose expression was perturbed by serum lipid traits were targets of statins, a category of drugs aiming at regulating the very same traits. Lipids are major modulators of CAD risk [56, 58] and established regulators of gene expression [59]. Hence, drugs targeting these downstream genes might modulate CAD risk, even though mediation analysis is warranted to support this hypothesis.

Combining results of DEG analysis and bi-directional TWMR allowed to decompose the observational correlation between whole blood gene expression and complex traits. This analysis showed that differentially expressed genes often reflect disease-induced changes in the transcriptome rather than disease-causing ones. Importantly, we observed that most of the correlation between gene expression and complex traits is due to confounders, which is plausible because age and sex are important determinants of both. The rest of the correlation can be almost entirely explained by the trait-to-gene expression causal effects. Indeed (just like single SNPs) individual genes’ expression has only minute contribution to complex traits, even if cumulatively they may contribute substantially. Diseases, however, represent a major burden for the organism, which can lead to drastic changes in the transcriptome program. In light of these considerations, it might be unsurprising that the correlation between a gene’s expression level and a complex trait is reflecting disease status rather a trait-to-expression link.

Like all methods, our approach has its limitations, which need to be considered when interpreting the results. First, our results are mainly focused on eQTLs and DEGs in whole blood. When conducting a tissue-specific analysis using GTEx data, we had considerably lower power to detect trait→gene associations because analysis of *trans*-eQTLs require much larger sample sizes than those collected by GTEx. However, because gene regulation is tissue-specific and many diseases manifest themselves only in certain tissues, the possibility to interrogate more tissues could unravel causative disease-gene links for genes not differentially expressed in blood. Furthermore, preliminary studies suggest that *trans-*eQTLs are particularly cell type-specific [60].

Then, TWMR and revTWMR results are difficult to compare because of the difference of power in the two approaches. One of the most important determinants of statistical power for MR is the sample size available for the outcome, thus revTWMR is less powered and prone to pick up mostly stronger effects. Still, another factor influencing power is the number and strength of instruments. Hence, TWMR results will be more accurate once larger eQTL data sets become available, which will in turn increase the number of testable genes (currently 16K). Finally, as every MR approach, revTWMR is at risk of violating the MR assumptions. In particular, horizontal pleiotropy and indirect effects of the instruments on the exposures can substantially bias causal effect estimates. RevTWMR assumes that the top GWAS SNPs have a direct effect on the phenotype. However, many SNPs show indirect or pleiotropic effects. We therefore protect our results from potential biases by excluding pleiotropic SNPs failing the heterogeneity test. Further gain in robustness should be obtained by including additional phenotypes (as exposures) through which instruments may act, as accounting for pleiotropy is a better approach than directly excluding violating instruments. Such multi-phenotype revTWMR approach will be possible only once genome-wide *trans*-eQTLs summary statistics will become available.

A very exciting perspective is that revTWMR can theoretically be extended to other types of *omics* data, e.g. integrating methylomics data, as alterations in DNA methylation are more often the consequence rather than the cause of diseases [61]. It would be interesting to apply revTWMR on protein levels as well (revPWMR), to gain further insights into the effects of complex traits on biomarkers but unfortunately, the sample size of proteomics datasets is still too small.

In conclusion, our bi-directional analysis disentangles causes and consequences of gene expression for complex traits and reveals that complex traits have more pronounced impact on gene expression than the reverse. Therefore, studies comparing gene expression levels of diseased and healthy subjects may still point to useful biomarkers of disease predisposition or severity, but interventions that restore levels of the biomarker to normal levels will not necessarily be disease modifying.

## Methods

### Reverse Transcriptome-wide Mendelian Randomization (revTWMR)

RevTWMR is a multi-instrument MR approach designed to estimate the causal effect of the phenotypes (exposure) on gene expression (outcome). For each gene, using inverse–variance weighted method for summary statistics [16], we define the joint causal effect of the phenotypes on the outcome as

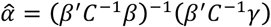

Here *β* is a *n-*vector that contains the effect size of *n* independent SNPs on the phenotype, derived from GWAS. *γ* is a vector of length *n* that contains the effect size, in *trans-*, of each SNP on the gene expression. *C* is the pair-wise LD matrix between the *n* SNPs.

As instrumental variables, we used independent (*r*^*2*^ < 0.01) significant (*P*_GWAS_ < 5×10^−08^) SNPs chosen among the 10K pre-selected trait-associated SNPs included in *trans*-eQTL dataset from eQTLGen Consortium (31,684 whole blood samples). As we are using only strongly independent SNPs, we use the identity matrix to approximate *C*.

The variance of α can be calculated approximately by the Delta method

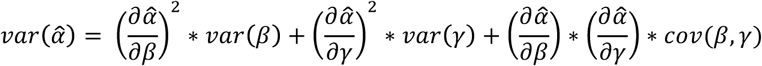

where *cov*(*β,γ*) is 0 if *β* and *γ* are estimated from independent samples (or if *β* and *γ* are independent). We defined the causal effect Z-statistic for gene *i* as 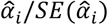, where 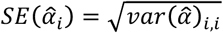.

We applied revTWMR across the human genome for causal association between a set of 12 phenotypes and the expression levels of 19,942 genes using summary statistics from GWAS and eQTLs studies. The analysed traits include: body mass index (BMI) [18], Crohn’s disease (CD) [17], educational attainment (EDU) [21], fasting glucose (FG) [19], high-density lipoprotein (HDL) [24], height [25], low-density lipoprotein (LDL) [24], rheumatoid arthritis (RA) [20], schizophrenia (SCZ) [22], total cholesterol (TC) [24], triglycerides (TG) [24] and waist-to-hip ratio adjusted for BMI (WHRadjBMI) [23]. All summary statistics (estimated univariate effect size and standard error) originate from the most recent meta-analyses and were downloaded from the publicly available NIH Genome-wide Repository of Associations Between SNPs and Phenotypes (https://grasp.nhlbi.nih.gov/). We only used SNPs on autosomal chromosomes and available in the UK10K reference panel, which allowed to estimate the LD among these SNPs and prune them. Strand ambiguous SNPs were removed.

### Heterogeneity test

The validity of all MR approaches, such as revTWMR, relies on three assumptions. The third assumption (no pleiotropy) is crucial as MR causal estimates will be biased if the genetic variants (IVs) have pleiotropic effects [50]. Hence, revTWMR assumes that all genetic variants used as instrumental variables affect the outcome only through gene expression and not through independent biological pathways. To test for the presence of pleiotropy, we used Cochran’s Q test [49, 51]. In brief, we tested whether there is a significant difference between the revTWMR-effect of an instrument (i.e. *αβ*_*i*_) and the estimated effect of that instrument on the gene expression (*γ*_*i*_). We defined

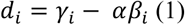

and its variance as

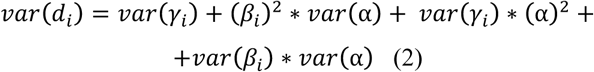

Next, we tested the deviation of each SNP using the following test statistic

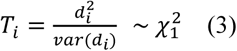

In case where *P* < 1×10^−4^, we removed the SNP with largest |*d*_*i*_| and then repeated the test.

### Transcriptome-wide Mendelian Randomization (TWMR)

In order to test the presence of a feedback loop of association, we ran TWMR [15] for all the significant revTWMR genes. The associations between the instrumental variables and the exposure (gene expression) and the outcome (complex traits) are estimated from the same studies used for revTWMR.

### Gene-based test

To compare GWAS and revTWMR results, we performed gene-based test for association summary statistics using PASCAL [46]. PASCAL assesses the total contribution of all SNP within close physical proximity to a given gene by combining SNP association Z-statistics into gene-based P-values while accounting for local LD structure.

### Replication cohorts

#### EGCUT

##### Study population

The Estonian Genome Center, University of Tartu (EGCUT) cohort denotes for the Estonian Biobank sample of more than 200,000 individuals, or about 20% of the Estonian adult population. All Biobank participants have been genotyped and linked to electronic health records (EHR) of the Health Insurance Fund, national registries and major hospitals. The EHR linkage captures the participants’ medical history together with demographics, lifestyle information and laboratory measurements; additional information is provided by self-completed questionnaires. Disease diagnoses are in the form of ICD-10 codes. RNA-seq data is available on 491 unrelated individuals. All Biobank participants have signed a broad informed consent to allow using their genetic and medical information for research purposes.

##### Whole-Blood-Transcriptome analysis

The preparation of RNA-seq data has been described in detail elsewhere [https://doi.org/10.1371/journal.pcbi.1005766] [62]. RNA-seq reads were trimmed of adapters together with low-quality leading and trailing bases using Trimmomatic (version 0.36) [63]. Additional quality control was performed with FastQC (version 0.11.2). The final set of reads were mapped to a human genome reference version GRCh37.p13 using STAR (version 2.4.2a) [64]. Sample mix-ups were tested and corrected for using MixupMapper [65]. Principal component analysis on RNA-seq read counts revealed a batch of outlying samples which was uncovered to be due to a technical problem in library preparation – affected samples were discarded. Data was normalized using weighted trimmed mean of M-values [66] and used as log2-transformed counts per million. To account for (hidden) batch effects, the sequencing batch date together with first gene expression principal components were used in all subsequent analyses.

#### InChianti

##### Study population

The InCHIANTI study is a population based sample that includes 298 individuals of <65 age and 1155 individuals of age ≥65 years. The study design and protocol have been described in detail previously [67]. The data collection started in September 1998 and was completed in March 2000. The INRCA Ethical Committee approved the entire study protocol.

##### Whole-Blood-Transcriptome analysis

Peripheral blood specimens were collected from 712 individuals using the PAXgene tube technology to preserve levels of mRNA transcripts. RNA was extracted from peripheral blood samples using the PAXgene Blood mRNA kit (Qiagen, Crawley, UK) according to the manufacturer’s instructions.

RNA was biotinylated and amplified using the Illumina® TotalPrep(tm) −96 RNA Amplification Kit and directly hybed with HumanHT-12_v3 Expression BeadChips that include 48 803 probes. Image data were collected on an Illumina iScan and analysed using Illumina GenomeStudio software. These experiments were performed as per the manufacturer’s instructions and as previously described [68]. Quality-control analysis of gene expression levels were previously described [69].

#### SHIP-Trend

##### Study population

The Study of Health in Pomerania (SHIP-Trend) is a longitudinal population-based cohort study in West Pomerania, a region in the northeast of Germany, assessing the prevalence and incidence of common population-relevant diseases and their risk factors. Baseline examinations for SHIP-Trend were carried out between 2008 and 2012, comprising 4,420 participants aged 20 to 81 years. Study design and sampling methods were previously described [70]. The medical ethics committee of the University of Greifswald approved the study protocol, and oral and written informed consents were obtained from each of the study participants.

##### Whole-Blood-Transcriptome analysis

Blood sample collection as well as RNA preparation were described in detail elsewhere [71]. Briefly, whole-blood samples of a subset of SHIP-TREND were collected from the participants after overnight fasting (≥10 hours) and stored in PAXgene Blood RNA Tubes (BD). Subsequently, RNA was prepared using the PAXgeneTM Blood miRNA Kit (QIAGEN, Hilden, Germany). Purity and concentration of RNA were determined using a NanoDrop ND-1000 UV-Vis Spectrophotometer (Thermo Scientific). To ensure a constantly high quality of the RNA preparations, all samples were analyzed using RNA 6000 Nano LabChips (Agilent Technologies, Germany) on a 2100 Bioanalyzer (Agilent Technologies, Germany) according to the manufacturer’s instructions. Samples exhibiting an RNA integrity number (RIN) less than seven were excluded from further analysis. The Illumina TotalPrep-96 RNA Amplification Kit (Ambion, Darmstadt, Germany) was used for reverse transcription of 500 ng RNA into double-stranded (ds) cDNA and subsequent synthesis of biotin-UTP-labeled antisense-cRNA using this cDNA as the template. Finally, in total 3,000 ng of cRNA were hybridized with a single array on the Illumina Human HT-12 v3 BeadChips, followed by washing and detection steps in accordance with the Illumina protocol. Processing of the SHIP-Trend RNA samples was performed at the Helmholtz Zentrum München. BeadChips were scanned using the Illumina Bead Array Reader. The Illumina software GenomeStudio V 2010.1 was used to read the generated raw data, for imputation of missing values and sample quality control. Subsequently, raw gene expression data were exported to the statistical environment R, version 2.14.2 (R Development Core Team 2011). Data were normalized using quantile normalization and log2-transformation using the lumi 2.8.0 package from the Bioconductor open source software (http://www.bioconductor.org/). Finally, 991 samples were available for gene expression analysis. Technical covariates used in all statistical models included RNA amplification batch, RNA quality (RIN), and sample storage time. The SHIP-Trend expression dataset is available at GEO (Gene Expression Omnibus) public repository under the accession GSE 36382: 991 samples were available for analysis.

### Phenotype-Gene expression correlation

To calculate the correlation between the phenotypes and the gene expression levels, we asked each cohort to run the following analysis. First, inverse normal transformation was applied to phenotypes and gene expression. Next, transformed phenotypes were adjusted only for sex, age and age^2^, while gene expression was also corrected for other known relevant covariates. Finally, Pearson’s correlation was calculated between the adjusted trait and the adjusted expression. Finally, correlations from single cohorts were combined using inverse variance meta-analysis, where weights are proportional to the squared standard error of the correlation estimates, as implemented in METAL [72].

### Observed and true correlation between gene expression and traits

The correlation between the effects estimated by revTWMR (*α*_*revTWMR*_) and the observational correlation (*corr(E, T*)) measured in the individual data from EGCUT, InChianti and SHIP-Trend was calculated using the Pearson’s correlation. As such correlation does not consider the error of the estimations, for the significant correlations we used the linear errors-in-variables models to compute the potential *true* correlation using the following equation

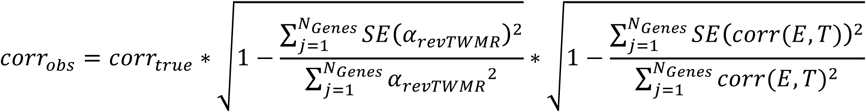

### Proportion of observational correlation explained by bi-directional causal effects

Let *E* and *T* denote the gene expression and the trait respectively. In addition there may exist a confounding factor *U* causally impacting both of them. We can express *E* and *T* as:

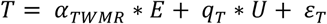

and

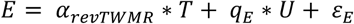

where *α*_*TWMR*_ and *α*_*revTWMR*_ are the causal effects of *E* on *T* and of *T* on *E* estimated by TWMR and revTWMR respectively; *q*_T_ and *q*_E_ are the causal effects of the confounders on *T* and *E*; and *ε* _*T*_ ∼*N*(0, *σ* _*T*_) and *ε* _*E*_ ∼*N*(0, *σ* _*E*_) represent uncorrelated errors. More specifically, *ε* _*T*_, *ε* _*E*_, and *U* are all independent of each other, because all dependence between *T* and *E* are due to bi-directional causal effects and the confounder *U*, the residual noises are independent of each other and of the confounder.

For simplicity, we assume that *E, T* and *U* have zero mean and unit variance, so that the correlation between *E* and *T* can be expressed as

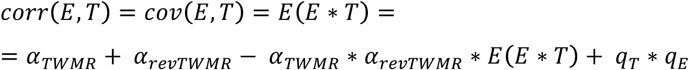

Equivalently,

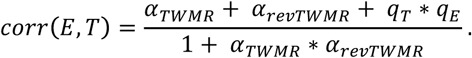

As we know the correlation, the bi-directional causal effects estimated by TWMR and revTWMR, we can estimate the contribution of the confounders (*q* _*T*_ * *q* _*E*_) to the observed correlation.

To avoid the recursive equations expressing the forward and reverse causal effects of E on T, we can substitute *T* into the equation for *E* and obtain

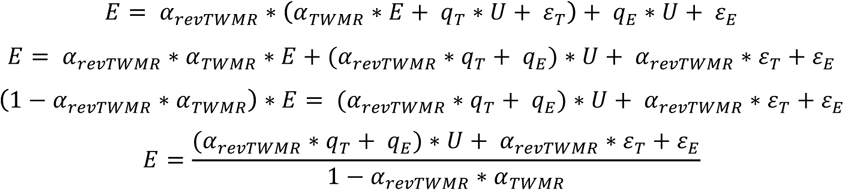

Similarly for *T*

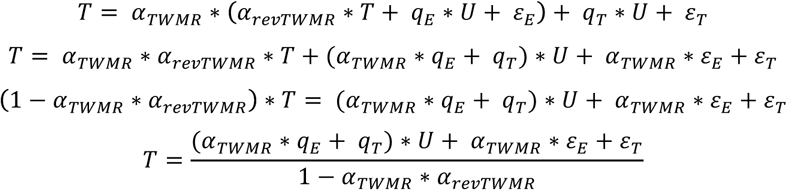

### GWAS hits trans-eQTL mapping in GTEx

Genotypes and gene expression quantifications from the GTEx project v8 dataset [54] were obtained via dbGaP accession number phs000424.v8.p1. This includes genotypes of 838 subjects, 85.3% of European American origin, 12.3% African American and 1.4% Asian American. The phased version of the genotype files was used and the genotypes for 2,177 out of 2,212 GWAS hits used as instrument variables in revTWMR were retrieved, matching for chromosome, position and reference/alternative allele, after conversion to GRCh38 coordinates using the UCSC liftOver tool [73]. Gene expression quantification (TPM values) from RNA-seq experiments across 49 tissues (for which genotype data is also available for >=70 individuals) processed and provided by the GTEx project v8 were also downloaded. These gene expression quantifications had been mapped to Gencode v26 [74] gene annotations on GRCh38 and normalised by TMM between samples (as implemented in edgeR), and inverse normal transform across samples. Moreover, only genes passing an expression threshold of >0.1 TPM in >=20% samples and >=6 reads in >=20% samples had been retained. The association between each of the 2,177 GWAS hits genotyped in GTEx v8 and each gene expression (20315 to 35007 genes per tissue, all gene types) across 49 tissues of the GTEx v8 was computed using QTLtools v1.3.1 trans function [75]. This consists of more than 2 billion association tests performed. For this, the --nominal option for calculating nominal p-values was used, as well as the --normal option, to enforce the gene expression phenotypes to match normal distributions N(0,1). To include all associations, no *cis* window filtering was applied. Moreover, covariates provided by GTEx v8 for each tissue were regressed out of each expression matrix to account for potential confounding factors, by using the -- covariate option on QTLtools. These included 15 to 60 PEER factors (depending on tissue sample size) [76], 5 Genotype PCA PCs as well as information about the sequencing platform, PCR usage, and the sex of the samples provided by GTEx v8.

## Data Availability

All the data are publicly available.
https://www.eqtlgen.org
https://grasp.nhlbi.nih.gov/FullResults.aspx.
No application for access nor registration was required to access the data from eQTLGen and GRASP

## Acknowledgments

This work was supported by grants from the Swiss National Science Foundation (310030-189147, 32473B-166450 to ZK and 31003A_182632 to AR) and Horizon2020 Twinning projects (ePerMed 692145 to AR). SHIP-Trend is part of the Community Medicine Research net of the University of Greifswald, Germany, which is funded by the Federal Ministry of Education and Research (grants no. 01ZZ9603, 01ZZ0103, and 01ZZ0403), the Ministry of Cultural Affairs as well as the Social Ministry of the Federal State of Mecklenburg-West Pomerania, and the network ‘Greifswald Approach to Individualized Medicine (GANI_MED)’ funded by the Federal Ministry of Education and Research (grant 03IS2061A). The University of Greifswald is a member of the Caché Campus program of the InterSystems GmbH. We would like to thank Liza Darrous and Ninon Mounier for their valuable feedback and comments on this manuscript.

## Author contributions

E.P. conceived and designed the study; E.P. and Z.K. contributed to the mathematical derivations of the research; E.P. performed statistical analyses; M.C.S. carried out drug target analyses; Z.K. supervised drug target analyses; K.L. has performed initial comparisons between gene expression-trait correlation and TWMR effects in the EGCUT cohort; C.A. and F.A.S. contributed with the biological interpretation of the results; E.P., A.R. and Z.K. drafted the manuscript; M.C.S. and C.A. contributed to the writing of specific sections; C.A. and F.A.S. revised the manuscript; K.L. performed statistical analyses on EGCUT cohort; A.M. oversaw the analysis in EGCUT; A.R.W. performed statistical analyses on InChianti cohort; S.B., T.T. and T.F. oversaw the analysis in InChianti; A.W. performed statistical analyses on SHIP-Trend cohort; U.V. and M.N. contributed to the data collection, quality control and study design of SHIP-Trend; A.T. oversaw the analysis in SHIP-Trend; D.M.R. performed *trans*-eQTLs analyses on GTEx dataset; O.D. designed and supervised *trans*-eQTLs analyses on GTEx dataset. All authors read the paper and contributed to its final form.

