## Supplementary Figures for "Differentially expressed genes reflect disease-induced rather than disease-causing changes in the transcriptome"

**Porcu et al.**

**Supplementary Figure 1**. ***Comparison between revTWMR and PASCAL results***. *For each gene we compared the* $-\log_{10}$*(P-value) calculated by revTWMR (x-axis) and PASCAL (y-axis).*

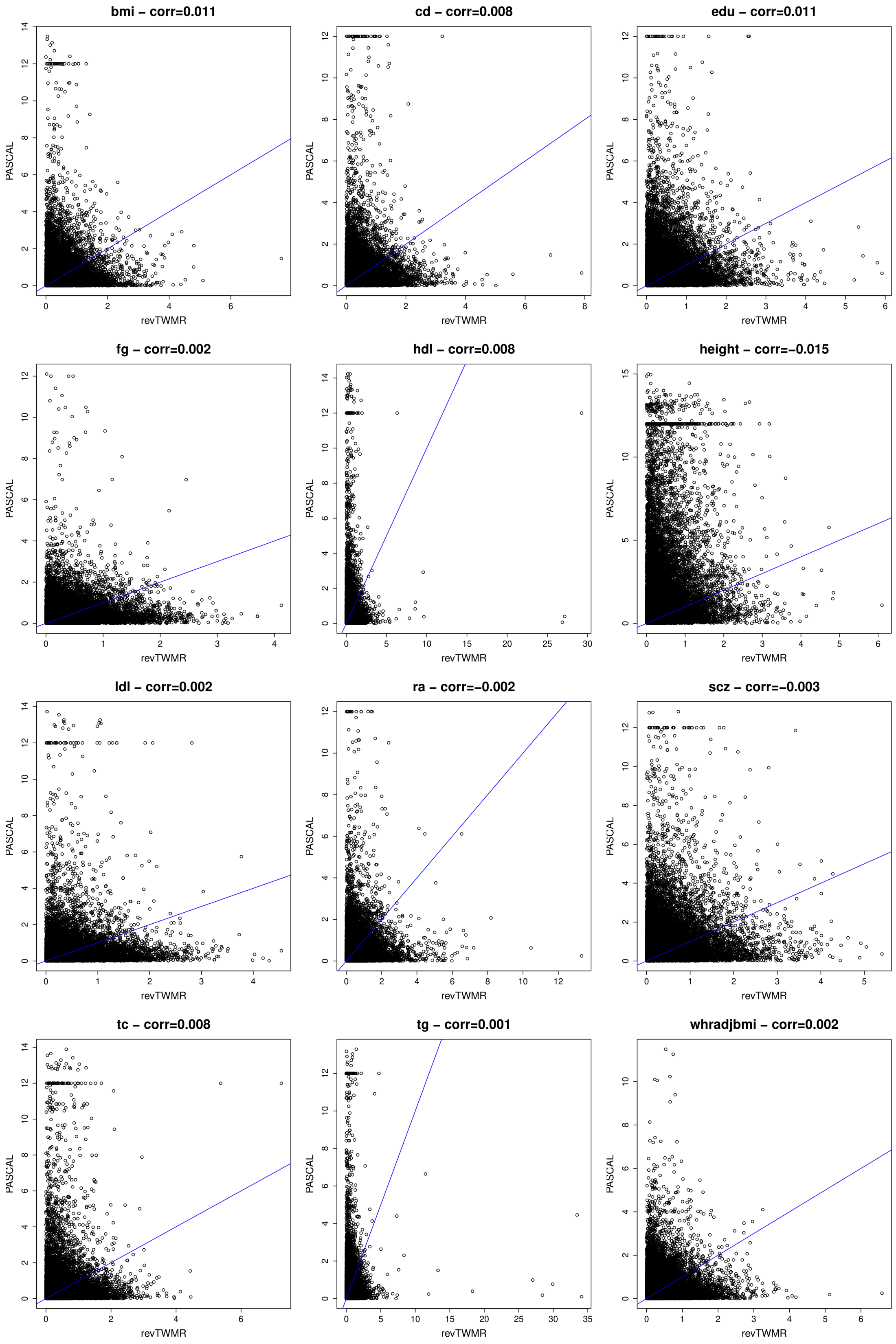

**Supplementary Figure 2**. ***Pleiotropic SNP leads to biased association for STX1B***. *For each SNP we plotted its effect on educational attainment (x-axis) and on the expression of STX1B (y-axis). The red and the green line represent the causal effect estimated by revTWMR before and after removing rs2456973 (red dot) respectively.*

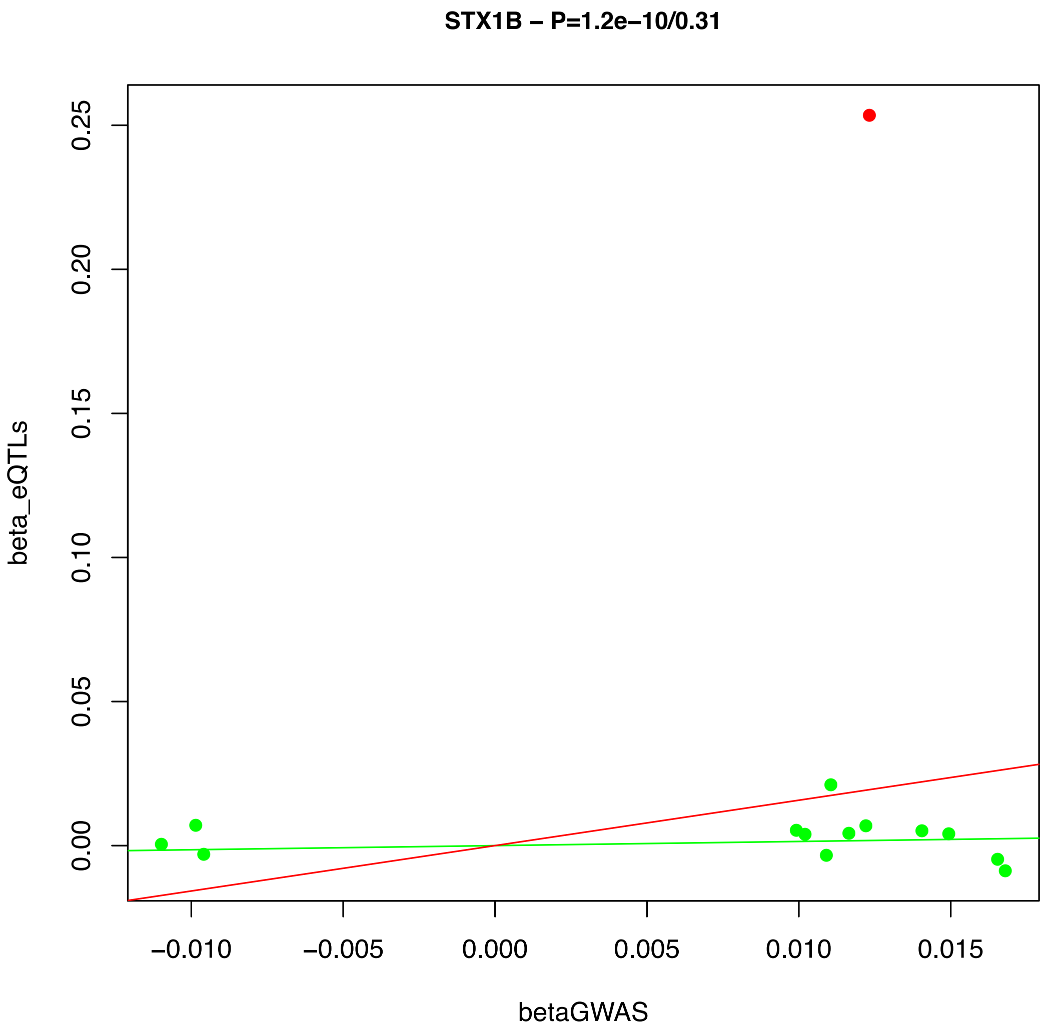

**Supplementary Figure 3**. ***Linear relationship between the perturbation correlation ($\hat{\rho}_P$) and the genetic correlation ($\hat{\rho}_G$) obtained from LD Score Regression***. *We selected the traits analyzed by our study and Bulik-Sullivan et al [1] and for each pair of traits we compared the two correlations. Gray dots represent non-significant trait pairs, blue dots represent trait pairs significant for both correlation and red and green ones correspond to pairs of traits significant only in $\hat{\rho}_G$ or $\hat{\rho}_P$, respectively. The dotted line represents the regression line*

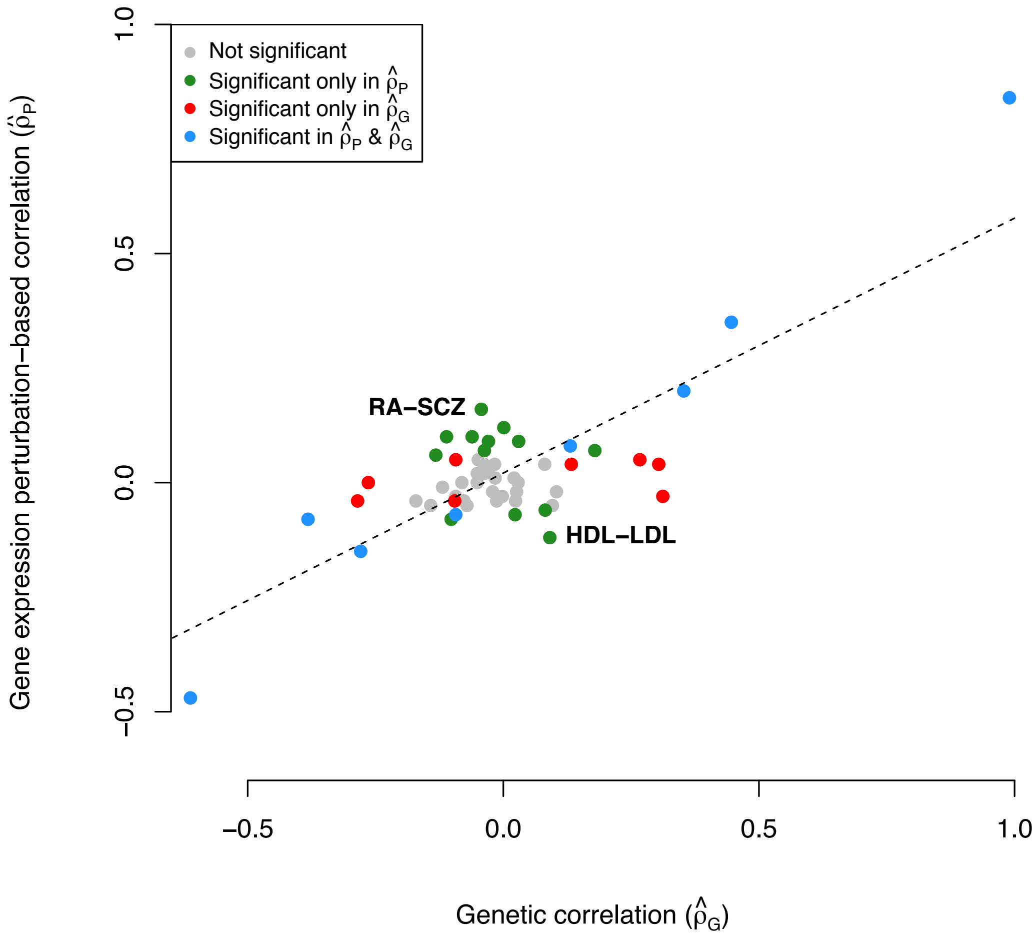

**Supplementary Figure 4**. ***Correlation of DEG results for LDL in the three independent cohort***. *For each gene we compared its expression-trait correlation in two cohorts. The blue line represents the identity line.*

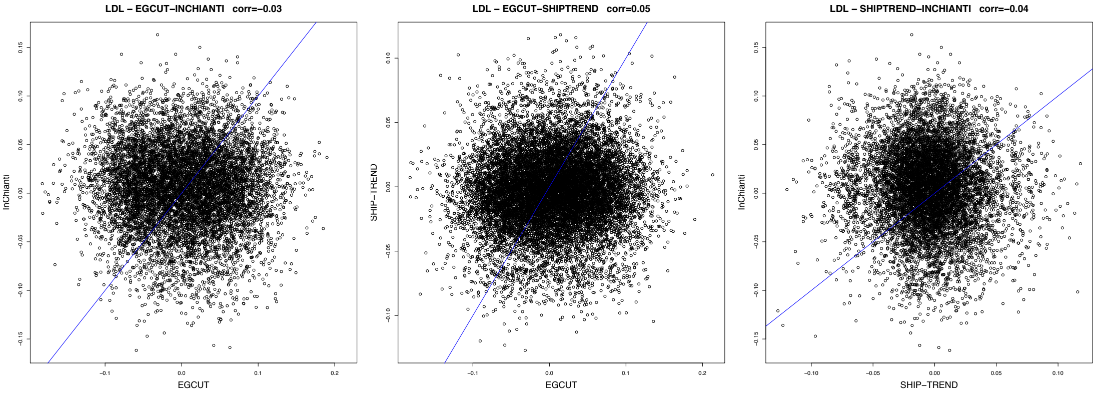
